# Elevated circulating monocytes and monocyte activation in pulmonary post-acute sequelae of SARS-CoV-2 infection

**DOI:** 10.1101/2022.11.19.22282543

**Authors:** Juwon Park, Logan S Dean, Boonyanudh Jiyarom, Louie Mar Gangcuangco, Parthav Shah, Thomas Awamura, Lauren L. Ching, Vivek R. Nerurkar, Dominic C. Chow, Fritzie Igno, Cecilia M Shikuma, Gehan Devendra

**Author notes:** **Correspondence:** Juwon Park, Ph.D., Assistant Professor, Department of Tropical Medicine, Medical Microbiology, and Pharmacology, John A. Burns School Medicine, University of Hawaii at Manoa, Honolulu, Hawaii, USA, 96813. **Co-correspondence:** Gehan Devendra, M.D., Assistant Professor, Division of Pulmonary and Critical Care, Queen’s Medical Center, Honolulu, HI, USA, 96813Honolulu, Hawaii, USA, 96813.

## Abstract

**Background:** Monocytes and macrophages play a pivotal role in inflammation during acute SARS-CoV-2 infection. However, their contribution to the development of post-acute sequelae of SARS-CoV-2 infection (PASC) are not fully elucidated.

**Methods:** A cross sectional study was conducted comparing plasma cytokine and monocyte levels among three groups: participants with pulmonary PASC (PPASC) with a reduced predicted diffusing capacity for carbon monoxide [DLCOc, <80%; (PG)]; fully recovered from SARS-CoV-2 with no residual symptoms (recovered group, RG); and negative for SARS-CoV-2 (negative group, NG). The expressions of cytokines were measured in plasma of study cohort by Luminex assay. The percentages and numbers of monocyte subsets (classical, intermediate, and non-classical monocytes) and monocyte activation (defined by CD169 expression) were analyzed using flow cytometry analysis of peripheral blood mononuclear cells.

**Results:** Plasma IL-1Ra levels were elevated but FGF levels were reduced in PG compared to NG. Circulating monocytes and three subsets were significantly higher in PG and RG compared to NG. PG and RG exhibited higher levels of CD169^+^ monocyte counts and higher CD169 expression was detected in intermediate and non-classical monocytes from RG and PG than that found in NG. Further correlation analysis with CD169^+^ monocyte subsets revealed that CD169^+^ intermediate monocytes negatively correlated with DLCOc%, and CD169^+^ non-classical monocytes positively correlated with IL-1α, IL-1β, MIP-1α, Eotaxin, and IFNγ.

**Conclusion:** This study present evidence that COVID convalescents exhibit monocyte alteration beyond the acute COVID-19 infection period even in convalescents with no residual symptoms. These data provide further rational for determining the role of monocyte subsets in PPASC pathogenesis.

## Introduction

It is estimated that one-third of patients infected with SARS-CoV-2 who develop coronavirus-19 (COVID-19) continue to experience residual symptoms, collectively referred to as ‘long-COVID’ or ‘post-acute sequelae of SARS-CoV-2 infection’ (PASC)^1^. PASC symptoms are highly heterogeneous with a wide range of presentations, including fatigue, dyspnea, sleep disorders, anxiety, and loss of memory and/or concentration. Among individuals with PASC, pulmonary complications, such as persistent dyspnea and chronic cough are common^2,3^. The pathophysiology of COVID-19 is complex and appears to involve multiple inflammatory and immunological pathways^4^. Studies have shown that COVID-19 patients display high systemic levels of cytokines and profound immune cell dysregulation that correlates with disease severity^5,6^.

Monocytes and macrophages are essential immune cells involved in host immunity and tissue homeostasis^7-9^. These cells also possess inflammatory^10^ and tissue-repairing capabilities and thus actively participate in all phases of the inflammatory response. Monocytes can be activated by infection and/or inflammatory conditions, leading to differentiation and polarization into macrophages with pro-inflammatory phenotypes^8,11,12^. High monocyte count and activated monocyte phenotype have been linked to various pathological conditions^12-16^. During SARS-CoV-2 infection, elevated peripheral monocyte levels and altered phenotype were observed in patients^13,14,17^. Analysis of circulating monocytes has shown to predict disease severity and mortality in COVID-19^18,19^. A comprehensive analysis of immune cells revealed long-term perturbations of innate and adaptive immune populations that persisted at least 6 months after SARS-CoV-2 infection^20^. COVID-19 convalescents with prolonged symptoms displayed highly activated myeloid cells, lacked naïve T and B cells, and exhibited elevated type I and III interferon levels^21^. A recent study found that intermediate (CD14^+^CD16^+^) and non-classical monocytes (CD14^dim^ CD16^+^) were significantly elevated in PASC patients. Furthermore, SARS-CoV-2 S1 protein was detected in non-classical monocytes, but not classical and intermediate monocytes in PASC patients suggesting that non-classical monocytes may contribute to inflammation in PASC^22^. Although our understanding of innate immunity underlying the pathophysiology of PASC is evolving, a detailed understanding of monocyte response in individuals with pulmonary PASC (PPASC) remain unclear. Given that blood monocytes provide a window into the systemic immune response, reflecting the risk of potential complications after recovery from acute infection, it is important to characterize these monocyte populations to gain insight into the role that monocyte dysregulation plays in PPASC.

In this study, we analyzed circulating monocytes and plasma cytokine expression in COVID-19 convalescents, comparing them to uninfected individuals. We further assessed the relationship between these parameters and quantitative measures of lung function. We found that COVID-19 convalescents, regardless of residual pulmonary symptoms, displayed increased monocyte levels and had an activated phenotype, defined as CD169^+^ cells. Moreover, the percentages and numbers of CD169^+^ monocyte subsets were associated with DLCOc% and proinflammatory cytokines, suggesting that alterations in monocyte subset activation may contribute to the development of chronic lung sequelae in individuals after resolution of COVID-19 infection.

## Methods

### Study cohort and selection of participants

This cross-sectional study investigated PASC complications among individuals living in Hawaii. Participants with PASC were recruited from the Post-COVID Recovery and Care Clinic of an academic tertiary care hospital (Queen’s Medical Center, Honolulu, Hawaii) between September 2020 and Mar 2021, prior to the detection of Omicron variants in Hawaii. Participants were grouped into the following: (1) individuals who reported persistent pulmonary symptoms (dyspnea, fatigue, cough, or shortness of breath) beyond 30 days after COVID-19 infection with reduced diffusion capacity for carbon monoxide (corrected for hemoglobin-DLCOc, <80%) by pulmonary function test (PPASC group [PG]; n=11); (2) individuals who have fully recovered from SARS-CoV-2 infection with no residual symptoms >30 days after acute infection (recovered group [RG]; n=10); and (3) individuals confirmed to have not contracted COVID-19 using negative SARS-CoV-2 antibody test (negative group [NG], n=10). The PG and RG groups had documented positive SARS-CoV-2 by polymerase chain reaction (PCR) and a replicated SARS-CoV-2 IgG antibody test. The study was approved by the Queens Medical Center Institutional Review Committee with the University of Hawaii IRB ceding authority (IRB#: RA-2020-053).

### Pulmonary function tests

Pulmonary function testing (PFT) was performed on individuals with PPASC. All PG group underwent PFTs (Vyaire) with the measurement of FVC, FEV1, TLC, and DLCOc% and interpreted in accordance with European Respiratory Society (ERS)/American Thoracic Society (ATS) guidelines^24^.

### Plasma and peripheral blood mononuclear cells (PBMC) isolation

Whole blood was collected from study participants in EDTA tubes (BD, Vacutainer) by venipuncture and processed based on a well-established method^25^. In brief, whole blood was centrifuged, plasma removed and cryopreserved at −80°C until downstream analysis. Remaining venous blood was diluted with an equal volume of PBS and layered on top of Ficoll-Paque Plus (GE Healthcare Biosciences, Piscataway, NJ) following the manufacturer’s protocol. PBMC were separated by centrifugation at 400 × g for 30 minutes at room temperature (RT). PBMC were collected from the buffy coat, red blood cells lysed, and then washed twice in PBS supplemented with 1% Fetal Bovine Serum (FBS). Cells were then counted, viability determined, and cryopreserved until further analysis.

### Multiplex Cytokine Analysis

Plasma was thawed and prepared following the manufacturer’s guidelines for each kit. All samples were run in duplicate in a single plate per panel. Biomarkers to assess inflammation (IFN-γ, IL-13, IL-1α, IL-8, IL-1β, IL-1rα, TNF-α), leukocyte chemotaxis (Eotaxin, MCP-1, MIP-3α, MIP-1α, RANTES/CCL5), and tissue remodeling/fibrosis (PDGF-AA, PDGF-AA/BB, FGF, VEGF, TGF-α) were measured using the R&D System™ Human XL Cytokine Discovery Premixed Kit. Data were acquired on a MAGPIX® Instrument (Luminex Corporation, Austin, TX). Data analysis was done using GraphPad Prism 9. Net median fluorescent intensity (MFI) was calculated (MFI value minus background value) and the average net MFI of duplicate samples was determined.

### Flow Cytometric Analysis

Cryopreserved peripheral blood mononuclear cells (PBMC) were thawed and washed twice in DPBS. Typically, 1-2×10^6^ cells were incubated LIVE/DEAD™ Fixable Yellow Dead Cell Stain (Invitrogen, 1:1000) at 4°C for 30 minutes, followed by addition of Human TruStain FcX (BioLegend, San Diego, CA, 1:200) in flow buffer (HBSS supplemented with 1% BSA) at RT for 15 minutes. Subsequently, cells were stained with the titrated fluorophore conjugated extracellular antibodies; CD45-BV711 (BD Biosciences, East Rutherford, NJ), CD11b-PE-Cy-7 (BioLegend, San Diego, CA), CD14-BV605 (BioLegend, San Diego, CA), CD16-BV421 (BioLegend, San Diego, CA), CD169-APC (Biolegend, San Diego, CA) at RT for 30 minutes and then washed twice with ice-cold flow buffer. Samples were then washed twice with ice-cold flow buffer and resuspended to 800 mL of flow buffer for acquisition. Samples were acquired with identical voltage settings on a LSR Fortessa (BD Sciences, East Rutherford, NJ) with approximately 1.0×10^9^ events collected per sample. 10 uL of AccuCheck Counting Beads (Life Technologies, Carlsbad, CA) were added prior to acquisition. Compensation beads (Invitrogen, Waltham, MA) were prepared for accurate compensation controls. Data was analyzed using FlowJo (Treestar, Ashland, OR) software and absolute cell counts were determined according to manufacturer protocols.

### Statistics

A cross sectional study comparing PG. RG, and NG participants was undertaken. Flow cytometry results between the groups were compared using Mann Whitney-U test. Patient characteristics between groups were compared using Chi-squared test, Fisher’s exact test, or Mann Whitney-U test, as appropriate. Correlation between monocyte subsets, CD169^+^ monocyte subsets, and inflammatory markers were analyzed using Spearman correlation. P-value < 0.05 was considered statistically significant for all tests. Statistical analyses were performed using IBM SPSS version 28 (Chicago, IL) and GraphPad Prism 9. (Graph Pad, San Diego, CA).

## Results

### Study cohort description

The median age of the participants was 53, 54, and 55 years for PG (n=11), RG (n=10), and NG (n=10), respectively. 18.2% of individuals with PPASC had respiratory history (asthma or COPD). Higher rates of hospital admission were seen among PG compared with RG (36% vs 0%, P = 0.015). No significant differences in pre-existing conditions, body mass index (BMI), and smoking prevalence between PG and RG was seen. Overall, 68.8% of the participants had received a SARS-CoV-2 vaccine at the time of enrollment. Individuals with PPASC experienced prolonged pulmonary complications including, dyspnea, fatigue, cough, and shortness of breath; 45.5% of them had at least two pulmonary symptoms. Those with PPASC reported symptoms lasting for a median duration of six months from post COVID-19 infection. All individuals from RG resolved symptoms within 4 weeks after disease onset and reported no symptoms at the time of sample collection. The baseline participant characteristics are displayed in **Table 1**.

**Table 1.** Characteristics of study participants.

|  | PG<br>(N=11) | RG<br>(N=10) | NG<br>(N=10) | P-value |
| --- | --- | --- | --- | --- |
| Age (years) | 53 [44, 58] | 54 [46.2, 59.2] | 55 [44, 58.2] | 0.953 |
| Male Gender | 8 (72.7%) | 6 (60%) | 6 (60%) | 0.778 |
| Months post-COVID infection | 6 [4, 12] | 5 [1.5, 9.5]* | -- | 0.515 |
| Body Mass Index (kg/m<sup>2</sup>) | 30.8 [24.3, 35.1] | 26.9 [22.0, 34.6] | No data | 0.676 |
| Hospital admission (n, %) | 4 (36.3%) | 0 | 0 | **0.015** |
| Race |  |  |  |  |
| White/Caucasian | 3 (27.3%) | 1 (10%) | 5 (50%) | 0.160 |
| Asian | 4 (36.4%) | 4 (40%) | 5 (50%) |  |
| Native Hawaiian/Pacific Islander | 3 (27.3%) | 2 (20%) | 0 |  |
| Other | 1 (9.0%) | 3 (30%) | 0 |  |
| Co-morbidities |  |  |  |  |
| Asthma | 1 (9.1%) | 1 (10%) | 1 (10%) | 0.997 |
| COPD | 1 (9.1%) | 0 | 0 | 0.391 |
| Hypertension | 3 (27.3%) | 2 (20%) | 1 (10%) | 0.605 |
| Diabetes | 2 (18.2%) | 1 (10%) | 1 (10%) | 0.809 |
| Smoking history (past/current) | 4 (36.4%) | 2 (20%) | 0 | 0.109 |
| Current alcohol use | 3 (27.3%) | 5 (50%) | 0 | **0.038** |

P-value calculated using Kruskal-Wallis test, Chi-square test, or Fisher's exact test.
\*1 missing data

### Soluble biomarkers levels in COVID-19 convalescents

To examine blood biomarkers associated with PPASC, we assessed 17 analytes in the plasma of participants from the NG, RG, and PG by Luminex assay and compared the plasma concentration of analytes among the groups. Analytes included cytokines associated with “cytokine storm”; IL-1α, IL-1β, IL-1Ra, IL-8, IL-13, and TNF-α. “leukocyte chemotaxis”; Eotaxin, MCP-1, MIP-3α, MIP-1α, Fractalkine, IP-10, MCP-3, RANTES), “tissue remodeling”; PDGF-AA, PDGF-AA/BB, FGF, VEGF, TGF-α. The IL-1Ra was elevated (2.2 fold higher) in the PG compared to NG, and its’ level trended higher in PG as compared to RG (**Figure 1A**). PDGF-AA, PDGF-AB, and TGFα were decreased in the RG, compared to the NG, and their levels tended to be lower in the PG (**Figure 1B-D**). FGF remained significantly lower in the RG and PG compared to NG (**Figure 1E**). There was no difference observed in 13 analytes among three groups (**Supplementary Figure 1**).

**Figure 1.**
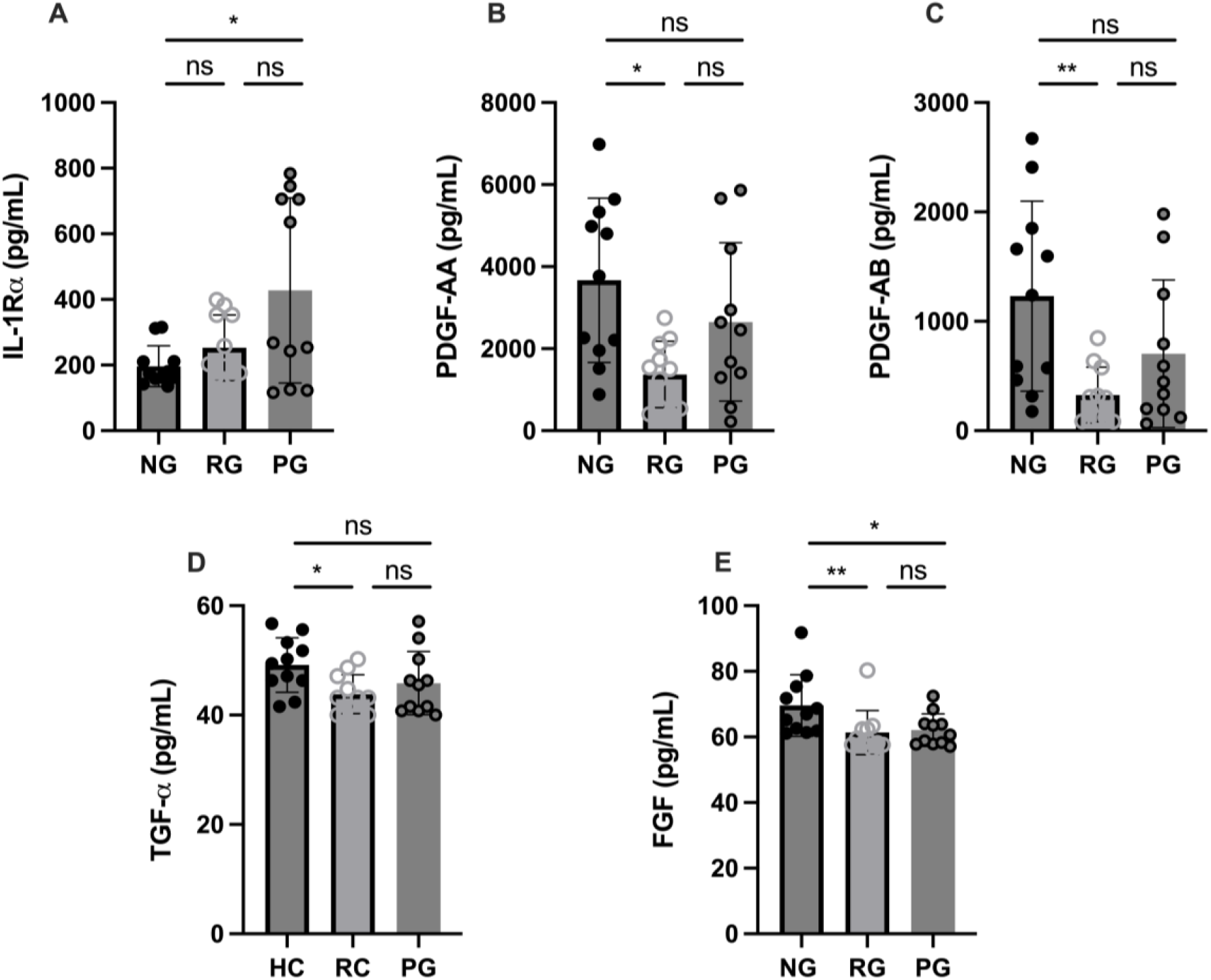
Plasma cytokine levels among groups. (**A**) IL-1Rα, (**B**) PDGF-AA, (**C**) PDGF-AB, (**D**) TGF-α, and **(E)** FGF levels were measured in plasma collected from NG, RG, and PG. The data are represented as the box and scatter plots with each circle representing a single individual. P values were calculated using the Mann–Whitney U-test. *p<0.05, **p<0.01, ns=non-significant.

### COVID-19 convalescents display elevated circulating monocytes

To determine the impact of long-term consequence of COVID-19 on blood monocytes and monocyte alteration in PPASC, PBMC cells isolated from NG (n=10), RG (n=10), and PG (n=11) cohorts were analyzed by flow cytometry. A representative gating strategy for identifying monocytes within the PBMC fractions from three groups was shown in Figure 2A. Monocyte subsets; classical (CD14^+^ CD16^-^), intermediate (CD14^+^ CD16^+^), or non-classical (CD14^dim^ CD16^+^) were defined by CD14 and CD16 surface levels within the monocyte (**Figure 2A**). We found both the percentages and numbers of total circulating monocytes were higher in the COVID-19 convalescents (PG and RG) than NG (**Figure 2B and C**). In comparison to NG, both the percentage and number of classical and intermediate monocytes were significantly increased in PG and RG (**Figure 3A, B, D, and E**). Also, the percentage and number of non-classical monocytes were significantly increased in PG, but only non-classical monocyte numbers were increased in RG, compared to NG (**Figure 3C and F**). However, we did not observe any significant difference between PG and RG in three monocyte subsets percentages and numbers (**Figure 3**). Altogether, these observations suggest that circulating monocyte levels remain elevated for several months after SARS-CoV2 infection, even convalescents who have no residual symptom.

**Figure 2.**
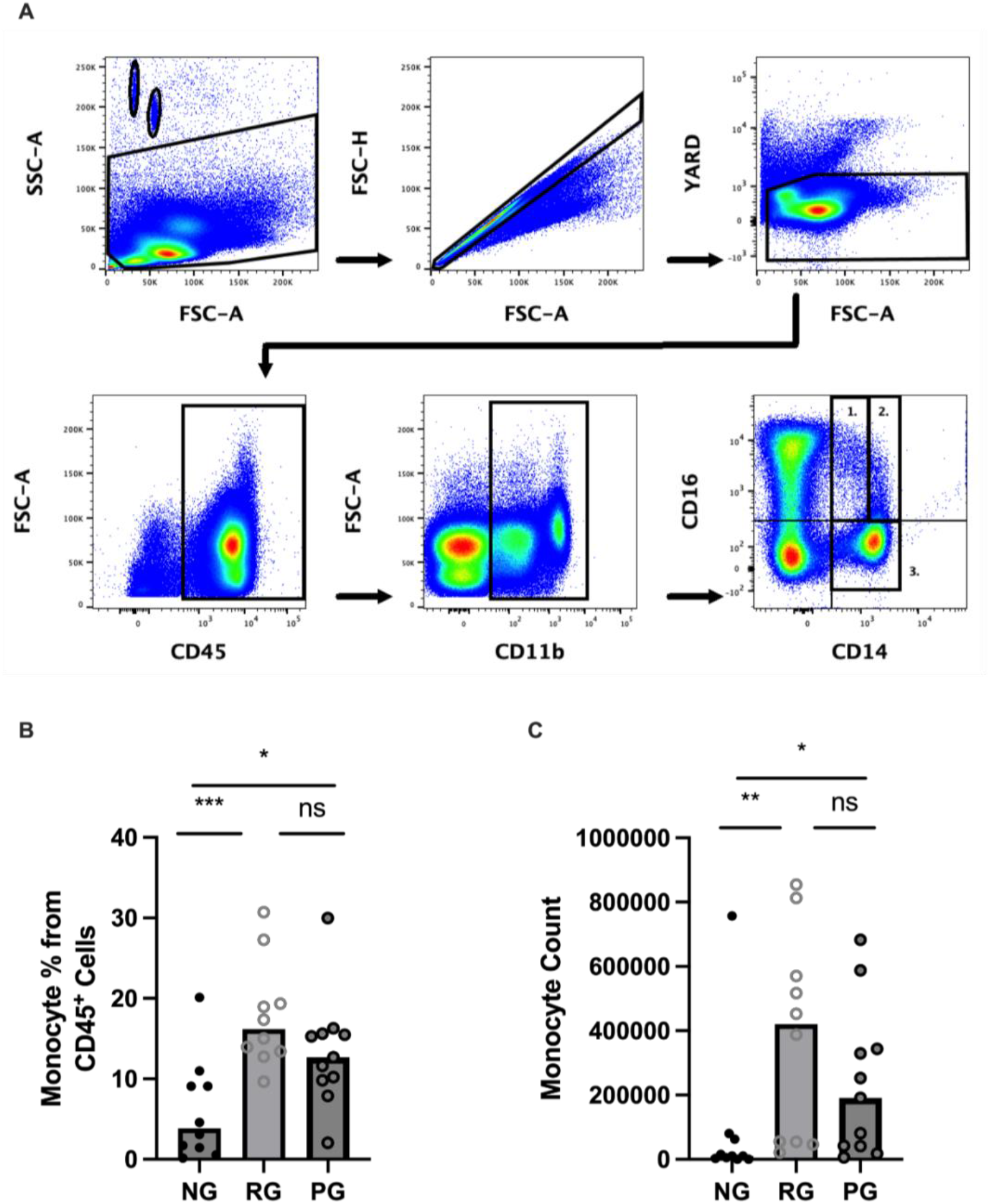
Comparison of circulating monocyte levels among groups. Representative flow cytometry gating strategy for identification of monocytes and monocyte subsets in PBMC from NG, RG and PG groups. Lymphocytes and monocytes were selected using CD45^+^ followed by gating for CD11b^+^ cells. (**A1**) non-classical (CD14^lo^/CD16^+^), (**A2**) intermediate (CD14^+^/CD16^+^, (**A3**) Classical (CD14^+^/CD16^-^) and (**B**) Total monocyte percentages as a proportion of total identified CD45^+^ cells in NG, RG, and PG groups. (**C**) Total monocyte counts in NG, RG, and PG groups. Mann-Whitney-U Test *p<0.05, **p<0.01, ***p<0.001, ns=non-significant.

**Figure 3.**
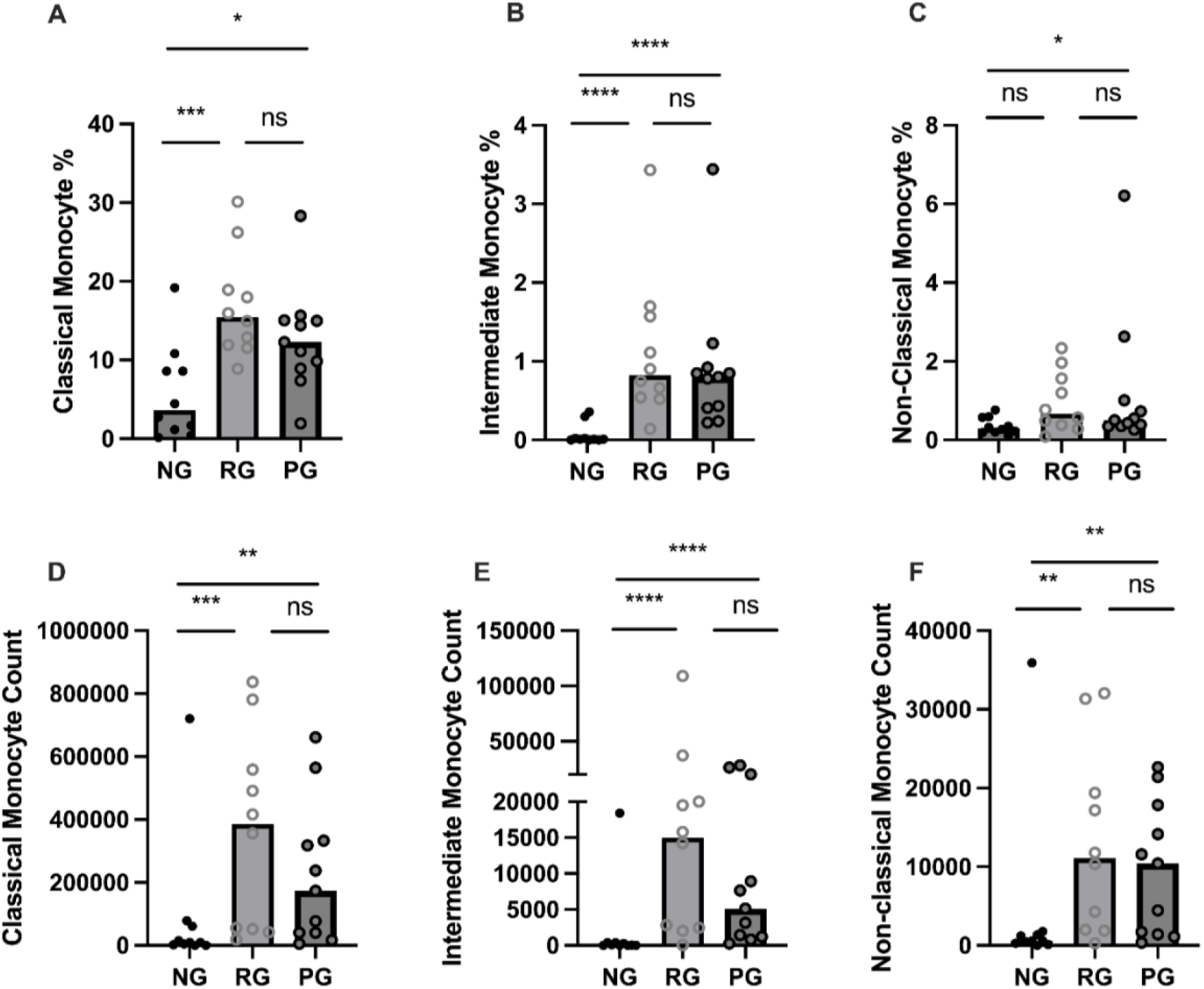
Comparison of circulating monocyte subsets among groups. (**A**) Classical monocyte percentage, (**B**) Intermediate monocyte percentage, (**C**) Non-Classical monocyte percentage, as a proportion of total identified CD45^+^ cells in NG, RG, and PG groups. **D**) Classical monocyte counts, (**E**) Intermediate monocyte counts, and (**F**) Non-Classical monocyte in NG, RG, and PG groups. Mann-Whitney-U Test *p<0.05, **p<0.01, ***p<0.001, ****p<0.0001, ns=non-significant.

### CD169^+^ monocytes in COVID-19 convalescents

CD169, a type I interferon-inducible receptor, is expressed on monocytes and macrophages^26-28^. CD169^+^ monocytes and macrophages have been thought to be important players in inflammatory response of inflammatory and autoimmune diseases^29-31^. Monocytes from COVID-19 patients were detected higher CD169 levels during acute SARS-CoV2 infection and monocyte CD169 was identified as a biomarker in early COVID-19 infection^27^. To further investigate difference in monocyte activation among the groups, we analyzed CD169 expression in monocytes. Baseline CD169 expression in monocytes from healthy donor was undetectable, so PMBC samples from quality control specimens were stimulated with increasing doses of IFNα (50, 100, and 200ng/mL) for discriminating monocytes expressing CD169 (**Supplementary Figure 2**). When monocytes were stratified based on CD169 expression, the percentage of CD169^+^ monocytes were significantly higher in the PG and RG than in the NG (**Figure 4A**). Also, CD169^+^ monocyte numbers were significantly increased in the PG and RG, compared to NG (**Figure 4B**). The mean fluorescence intensity (MFI) of CD169 on classical monocytes did not differ among the groups. However, CD169 MFI of intermediate and non-classical monocytes in PG and RG was significantly higher than those in NG (**Figure 4C**). Interestingly, when the percentage of CD169^+^ cells was examined in the three groups in each respective monocyte population, no difference was observed in classical monocytes (**Figure 4D**). Significant increases in CD169^+^ percentages were observed in intermediate and non-classical monocytes, only between RG and NG, with no difference between PG and NG, or PG and RG observed (**Figure 4E and F**). These data indicate that circulating monocytes from COVID-19 convalescents remain increased and display a higher CD169 expression.

**Figure 4.**
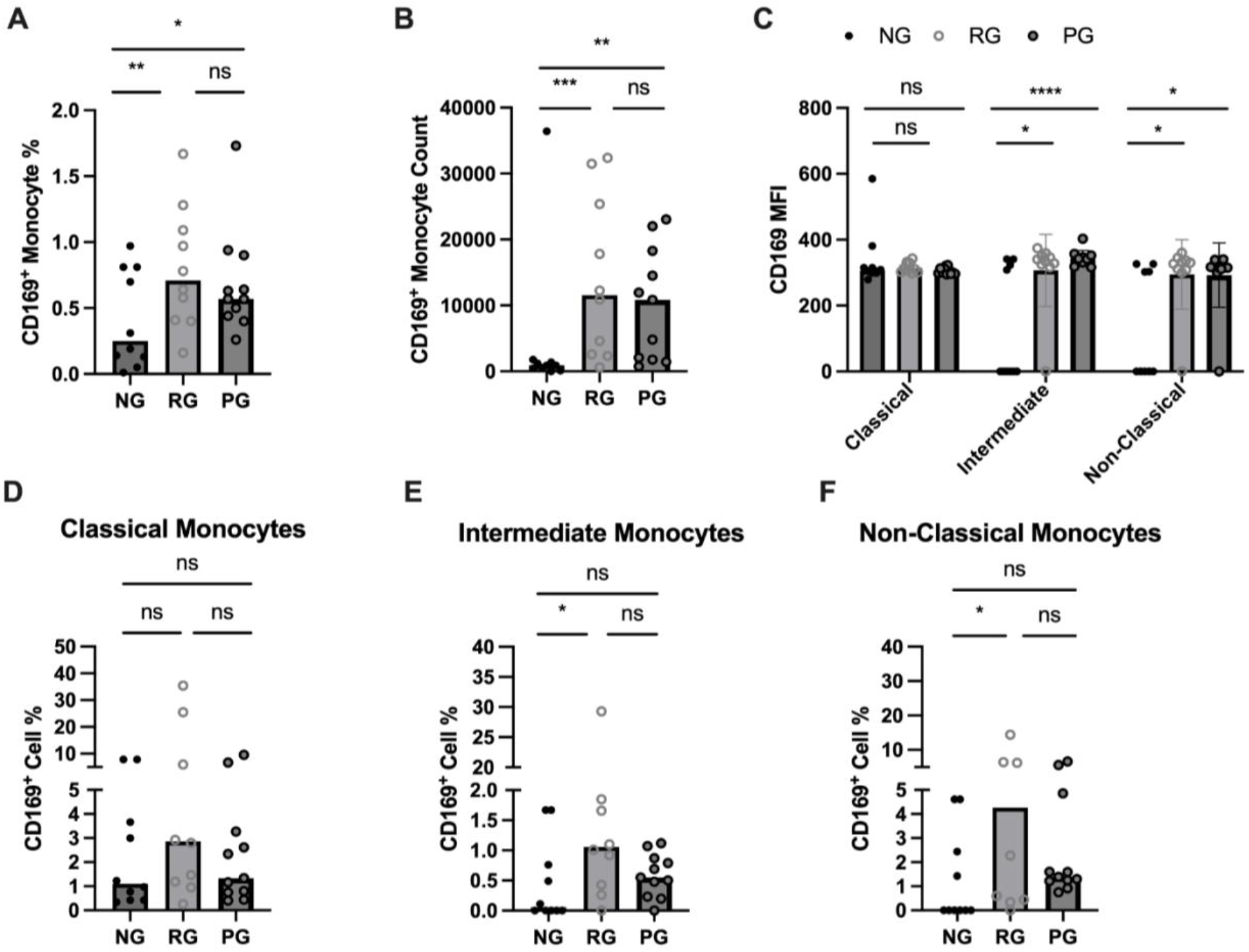
Characterization of circulating CD169^+^ monocytes in NG, RG, and PG groups. (**A**) Total CD169^+^ monocyte percentage from CD45^+^ cells, (**B**) CD169^+^ monocyte count (**C**) MFI of CD169 on monocyte subsets in NG, RG, and PG. The percentage of CD169^+^ cells identified in (**D**) classical monocytes, (**E**) intermediate monocytes, (**F**) non-classical monocytes within NG, RG, and PG groups. Mann-Whitney-U Test *p<0.05, **p<0.01, ***p<0.001, ****p<0.0001, ns=non-significant.

### The relationship between CD169^+^ monocytes and lung function in PPASC

PG participants had PFTs performed during their period of prolonged respiratory symptoms. 18.2% of participants reported pre-existing pulmonary conditions prior to infection with SARS-CoV-2 (**Table 1**). We explored the relationship between the percentages and counts of CD169^+^ monocyte with DLCOc%. We found negative correlations between CD169^+^ monocytes and CD169^+^ intermediate monocytes percentages and CD169^+^ intermediate monocyte counts (r= -0.758; P=0.009, r= -0.71; P=0.02, r= -0.69; P=0.051, respectively) and DLCOc% in PG (**Figure 5A-C**). Taken together, these results suggested that elevated CD169 expression in monocytes may serve as biomarker for determining lung function in PPASC.

**Figure 5.**
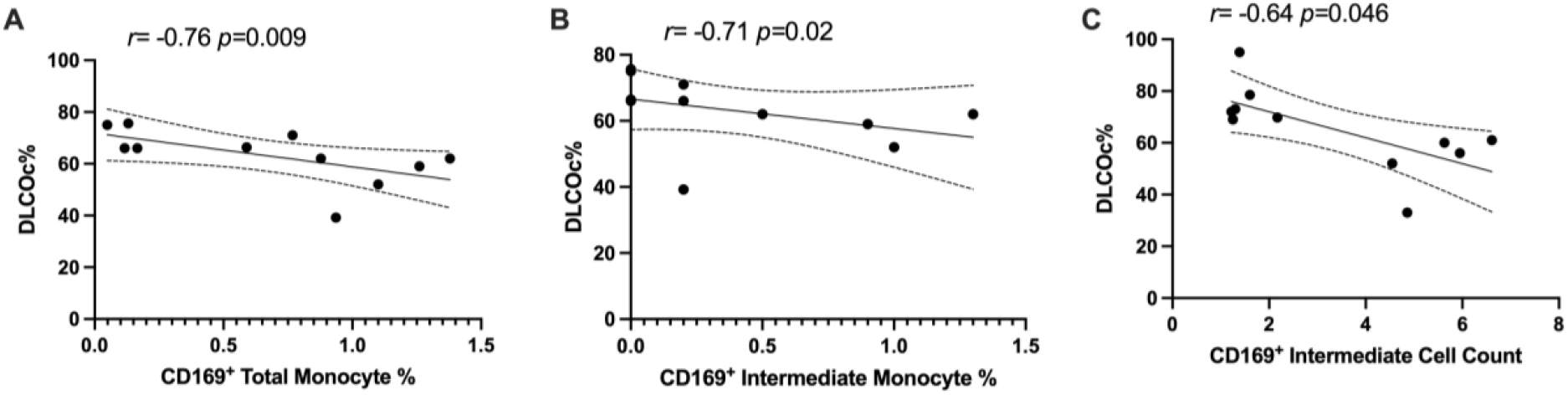
CD169^+^ monocytes were associated with DLCOc% in PG. Spearman correlation between DLCOc% and (**A**) percentage of CD169^+^ total monocytes, (**B**) percentage of CD169^+^ intermediate monocytes, and (**C**) CD169^+^ intermediate monocyte count.

### Monocyte relationship with systemic levels of cytokines in PPASC

In order to determine whether the changes in monocytes correlated with cytokine expression in PPASC, we performed spearman rank correlation to assess associations between cytokines and monocyte subset counts and percentages. In table 2, monocyte parameters (monocyte subsets and CD169^+^ total monocyte and monocyte subsets) showed a positive correlation with cytokines (VEGF, IL-8, IL-1α, IL-1β, PDGF-AA, PDGF-AB/BB, Eotaxin, MIP-1α, MCP-1, and IFN-γ).

**Table 2.** Spearman correlations of circulating molecules and monocyte populations in PG.

| Parameter | Analyte | R-value | P-value |
| --- | --- | --- | --- |
| Total CD169<sup>+</sup> Monocyte Count | VEGF | 0.84 | 0.002 |
| Intermediate Monocyte % of CD45<sup>+</sup> Cells | MIP-1α | 0.611 | 0.05 |
| | IL-1α | 0.709 | 0.017 |
| | IL-1β | 0.8 | 0.005 |
|  | PDGF-AA | 0.764 | 0.009 |
|  | PDGF-AB/BB | 0.782 | 0.006 |
|  | FGF | -0.627 | 0.044 |
| Intermediate Monocyte Count | Eotaxin | 0.627 | 0.044 |
| | MIP-1α | 0.63 | 0.042 |
|  | VEGF | 0.736 | 0.013 |
| Non-classical monocyte % of CD45<sup>+</sup> cells | IL-8 | 0.747 | 0.04 |
|  | MCP-1 | -0.709 | 0.018 |
| Non-classical Monocyte Count | VEGF | 0.836 | 0.002 |
| CD169<sup>+</sup> % of Intermediate Monocytes | MCP-1 | 0.691 | 0.022 |
| CD169<sup>+</sup> Non-classical Monocyte % of CD45<sup>+</sup> cells | IL-1α | 0.618 | 0.047 |
| | IL-1β | 0.745 | 0.011 |
| CD169<sup>+</sup> Non-classical Monocyte Count | IFN-γ | 0.636 | 0.04 |
|  | Eotaxin | 0.655 | 0.034 |
|  | PDGF-AA | 0.618 | 0.048 |
| | IL-1 $\alpha$ | 0.68 | 0.024 |
| | MIP-1α | 0.783 | 0.006 |

VEGF levels were positively associated with CD169^+^ total monocyte counts and non-classical monocyte percentages. The percentage of intermediate monocytes was positively associated with IL-1α, IL-1β, MIP-1α, PDGF-AA, and PDGF-AB/BB cytokines, but negatively associated with FGF. Eotaxin, MIP-1α, and VEGF were positively associated with intermediate monocyte counts, while MCP-1 was associated with the CD169^+^ percentage of intermediate monocytes. IL-1α and IL-1β were positively correlated with the percentage of CD169^+^ non-classical monocytes. IL-1α, MIP-1α, IFN-γ, Eotaxin, and PDFG-AA were positively correlated with CD169^+^ non-classical monocyte counts (**Table 2 and Figure 6)**. Altogether, these results suggested that monocyte subsets and the cells with CD169 upregulation were associated with a proinflammatory cytokine environment in PPASC.

**Figure 6.**
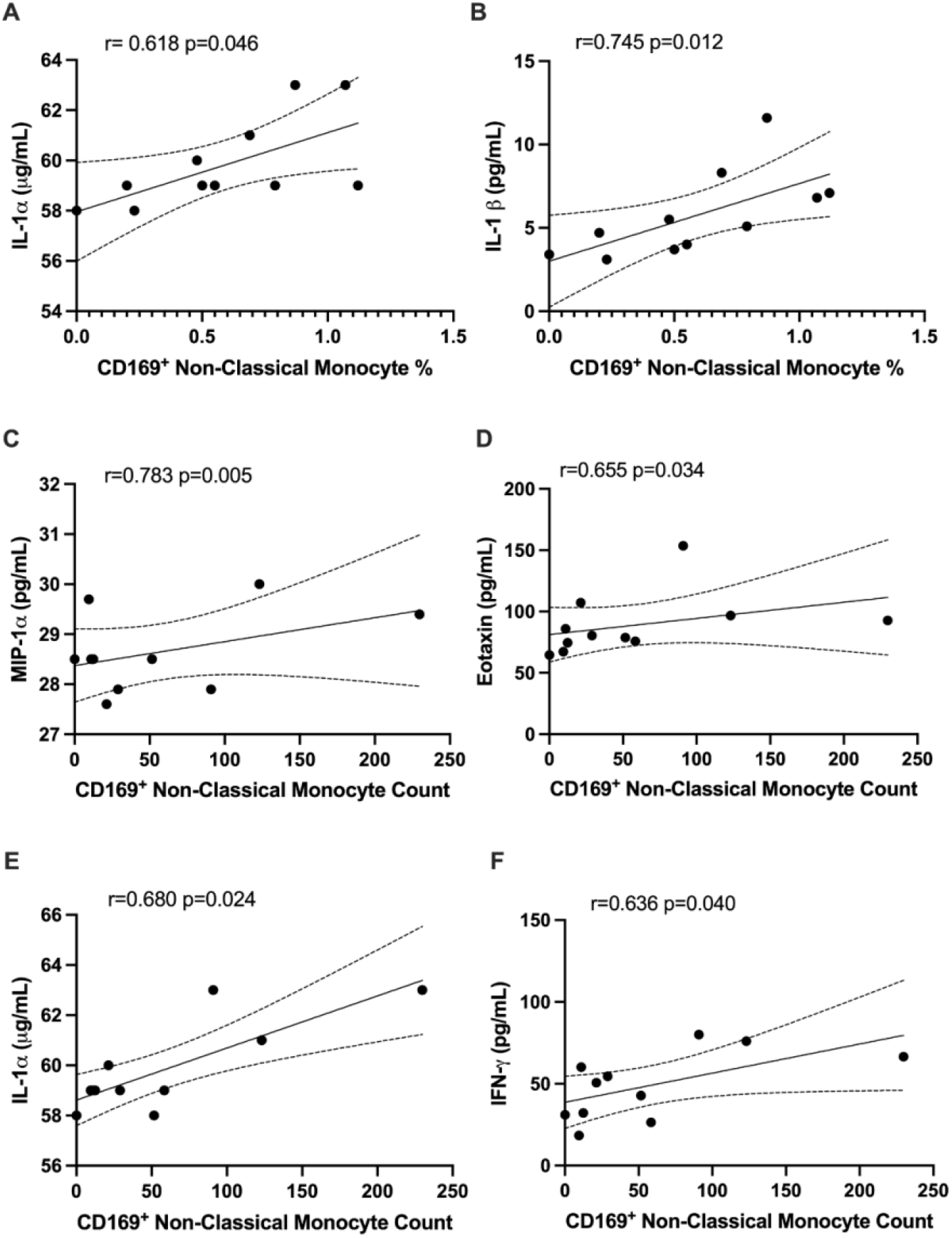
CD169^+^ non-classical monocytes were associated with IL-1α, IL-1β, MIP-1α, Eotaxin, and IFNγ in PG. Spearman correlation between the percentage of non-classical CD169^+^ monocytes and (**A**) IL-1α and (**B**) IL-1β. Spearman correlation between CD169+ non-classical monocyte count and (**C**) MIP-1α, (**D**) Eotaxin, (**E**) IL-1α, and (**F**) IFNγ.

## Discussion

In this study, we observed that COVID-19 convalescents with pulmonary PASC display altered circulating monocyte levels and activation, which may last several months after infection. Interestingly, monocyte alterations were also observed in individuals whose symptoms have resolved completely. These findings highlight that COVID-19 convalescents exhibit monocyte dysregulation beyond the resolution of initial infection, although larger studies are needed to validate this observation.

Few studies have detailed the immune profiles in individuals with PASC despite accumulating evidence increasingly indicating that immune cells contribute to persistent symptoms. It is still largely unknown if immune cells contribute to the immunopathology of PPASC. This is the first study, to our knowledge, to evaluate the associations of monocyte levels with lung function in a community-based cohort. In this study, we selected individuals with PPASC through a primary questionnaire and secondary evaluation of pulmonary function. This approach clarified the presence of pulmonary symptoms and may not discern individuals who had their symptoms overestimated based on the questionnaire. Studies demonstrate that patients with a mild case of COVID-19 frequently develop residual symptoms. Also, pulmonary symptoms were presented without impairment of lung function or cardiopulmonary exercise test among COVID-19 survivors^32,33^. However, monitoring of PFTs in severe COVID-19 survivors with lung abnormalities post discharge demonstrated significant pulmonary sequelae^34,35^. This study demonstrates that hospitalized COVID-19 survivors were more likely to have persistent pulmonary PASC symptoms compared with those without hospitalization. Hospitalized COVID-19 survivors tended to have reduced DLCOc% (61.98%) compared to non-hospitalized COVID-19 survivors (66.89%). A correlation analysis of CD169^+^ monocyte subsets and DLCOc% suggested that alteration of activated monocyte subsets may impact pulmonary function in COVID-19 convalescents. Nonetheless, we cannot exclude the possibility that this association may reflect other residual symptoms because a nonnegligible proportion of PPASC individuals also experienced other symptoms.

The cytokine profile revealed no changes in major inflammatory cytokines between NG, RG, and PG. Similar observations in COVID-19 convalescents have been reported in another cross-sectional study. IL-1α, IL-1β, IL-8, IFN-γ, VEGF-A, and TNF-α had returned to normal levels 6 months after recovery, but IL-1Rα was still elevated in COVID-19 convalescents, compared to healthy controls^19^. Another study showed that COVID-19 convalescents at 4 months post infection had higher levels of IFN-β, IFN-λ1, CXCL9, CXCL10, IL-8 and sTIM-3, regardless of symptoms compared to uninfected controls. IFN-β and IFN-λ1 still remained elevated in COVID-19 convalescents with PASC but others’ expressions were reduced at month 8, compared to month 4^21^. Oher studies investigating immune features of COVID-19 convalescent trends observed elevated levels of IL-6 and IL-1β^19,36,37^ in individuals with PASC. Interestingly, using published scRNA-sequencing datasets generated from severe COVID-19 patients demonstrated increased transcript reads of IL-1β and TNF-α from bronchoalveolar lavage fluids (BALF) macrophages^37^, supporting their hypothesis that proinflammatory reprogramming of lung macrophages or precursor monocytes, may drive prolonged and exacerbated PASC symptomology.

Some discrepancies in the reported cytokine levels from PASC studies continue to generate questions regarding the importance of a heterogeneous multisystemic condition. One might speculate the varying windows of sample collection post-infection between various studies alters the detectable cytokine profiles. Another possibility is that COVID-19 convalescents are not classified into symptomatic and asymptomatic. In this case, less systemic inflammation is represented, but respiratory environment may show distinct proinflammatory conditions in individuals with PPASC. Further analysis of cellular composition and cytokines in blood and BALF from individuals with PPASC over a longitudinal period is required to understand the dynamic features of respiratory and systemic immunity in PPASC during disease progression and resolution.

While no difference in major inflammatory cytokines between the three groups was observed, correlations of CD169^+^ monocyte subsets with cytokines suggested that specific activated monocyte subsets produce high levels of proinflammatory cytokines in PPASC. Correlations of D-dimer with CD169^+^ non-classical monocytes observed herein were corroborated by the findings of Pandori et al. (2022) in a cohort of individuals hospitalized for COVID-19^38^. Interestingly, their cohort did not display the increases in total monocyte populations in their hospitalized group but displayed decreases in non-classical monocyte percentages and steady levels of classical and intermediate monocyte percentages from total CD45^+^ cells in participants hospitalized for COVID-19 up to 90 days following admission. These trends suggest that decreased monocyte proportions are present during hospitalization from COVID-19, but COVID-19 convalescents demonstrated elevated monocyte levels, potentially in a dysregulated nature^19,21,22^. Further questions are raised as to whether monocyte subsets represent a key inflammatory driver of PPASC pathogenesis and are therefore a suitable biomarker for PPASC prognosis prediction and prognosis.

The COVID-19 pandemic has disproportionately impacted racial/ ethnic minority groups, including Black and Latinos^39,40^. In Hawaii. the COVID-19 pandemic has also exposed the stark health disparities experienced by Native Hawaiians and Other Pacific Islanders (NHOPI) and Filipinos. The NHOPI case rate and case fatality rate are 3-5 times and 4-10 times higher compared to Caucasian/White, respectively^41^. Among Filipinos, the hospitalization rates of COVID-19 have been reported to be 1.6 times higher than Whites^42^. In this study, participants composed primarily of non-White individuals in Hawaii, but we did not find any differences in the proportion PPASC versus non-PPASC patients by ethnicity likely due to the small sample size of the study.

This study was limited by the small sample size and was therefore not adequately powered for multivariable analyses. The length of post COVID-19 infection was variable,1 to 10 months post-infection. This variability in time of sample collection may have influenced monocyte population characteristics. Larger studies are warranted to further elucidate the role of circulating monocytes in PPASC. Also, a longitudinal evaluation of monocytes dynamics and the phenotypic changes after COVID-19 infection should be carried out to determine whether monocytes dysfunction is associated with the clinical outcome of respiratory failure. It would also be of clinical interest to track the perturbations of monocyte populations in relation to acute infection, COVID-19 disease context, and then into PPASC development or recovery.

In summary, these data indicate that systemic monocyte alteration continues COVID-19 convalescents with pulmonary symptoms, which is also found in COVID-19 convalescents with no residual symptoms. Also, COVID-19 convalescents exhibit activated monocyte phenotypes, denoted by CD169 expression, and that this activated phenotype is associated with poor lung function and increased proinflammatory cytokines. The drivers of PPASC pathogenesis require further investigation, but possibilities include high circulating monocyte levels, increased CD169^+^ intermediate and non-classical monocytes, and IL-1Rα expression. These observations will aid in informing the ongoing decipherment of the immunopathology that contributes to PPASC development, COVID-19 recovery, and subsequent therapeutic interventions.

## Data Availability

All data produced in the present study are available upon reasonable request to the authors.

---

**Supplemental Figure 1.**
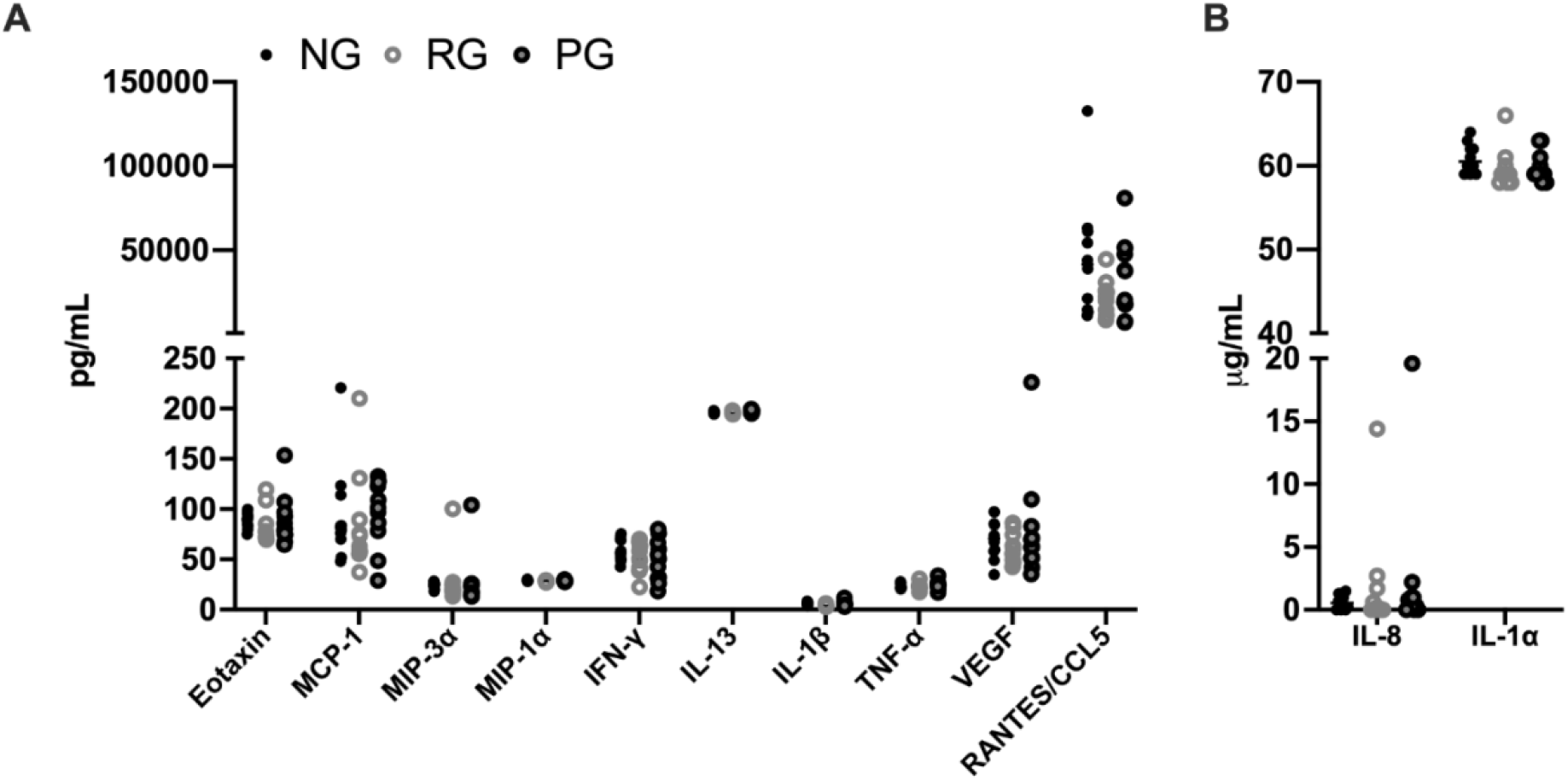
Levels of plasma cytokine in NG, RG, and PG. (**A**) No differences in Eotaxin, MCP-1, MIP-3α, MIP-1a, IFNγ, IL-13, IL-1β, TNFα, VEGF, RANTES/CCL5, (**B**) IL-8 and IL-1α.

**Supplemental Figure 2.**
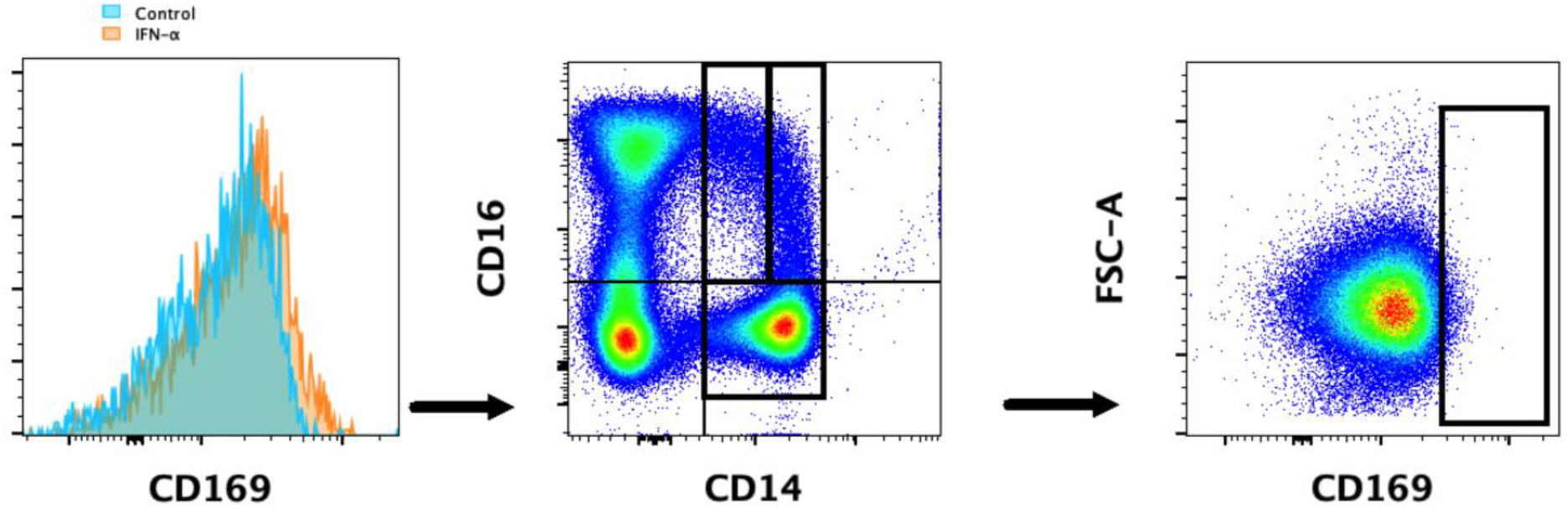
Histogram demonstrating the CD169 signal shift in response to 100 ng/mL IFN-α stimulation in quality control PBMC samples. This shift was used to develop gating for CD169^+^ monocyte populations

## References

1. Groff D, Sun A, Ssentongo AE, et al. Short-term and Long-term Rates of Postacute Sequelae of SARS-CoV-2 Infection: A Systematic Review. JAMA Netw Open 2021; 4(10): e2128568.

2. Jiang DH, Roy DJ, Gu BJ, Hassett LC, McCoy RG. Postacute Sequelae of Severe Acute Respiratory Syndrome Coronavirus 2 Infection: A State-of-the-Art Review. JACC Basic Transl Sci 2021; 6(9): 796–811.

3. Montani D, Savale L, Noel N, et al. Post-acute COVID-19 syndrome. Eur Respir Rev 2022; 31(163).

4. Merad M, Blish CA, Sallusto F, Iwasaki A. The immunology and immunopathology of COVID-19. Science 2022; 375(6585): 1122–7.

5. Terpos E, Ntanasis-Stathopoulos I, Elalamy I, et al. Hematological findings and complications of COVID-19. Am J Hematol 2020; 95(7): 834–47.

6. Giamarellos-Bourboulis EJ, Netea MG, Rovina N, et al. Complex Immune Dysregulation in COVID-19 Patients with Severe Respiratory Failure. Cell Host Microbe 2020; 27(6): 992–1000 e3.

7. Bassler K, Schulte-Schrepping J, Warnat-Herresthal S, Aschenbrenner AC, Schultze JL. The Myeloid Cell Compartment-Cell by Cell. Annu Rev Immunol 2019; 37: 269–93.

8. Guilliams M, Mildner A, Yona S. Developmental and Functional Heterogeneity of Monocytes. Immunity 2018; 49(4): 595–613.

9. Knoll R, Schultze JL, Schulte-Schrepping J. Monocytes and Macrophages in COVID-19. Front Immunol 2021; 12: 720109.

10. Zhou Y, Fu B, Zheng X, et al. Pathogenic T-cells and inflammatory monocytes incite inflammatory storms in severe COVID-19 patients. Natl Sci Rev 2020; 7(6): 998–1002.

11. Schulte-Schrepping J, Reusch N, Paclik D, et al. Severe COVID-19 Is Marked by a Dysregulated Myeloid Cell Compartment. Cell 2020; 182(6): 1419–40 e23.

12. Liao M, Liu Y, Yuan J, et al. Single-cell landscape of bronchoalveolar immune cells in patients with COVID-19. Nat Med 2020; 26(6): 842–4.

13. Liu N, Jiang C, Cai P, et al. Single-cell analysis of COVID-19, sepsis, and HIV infection reveals hyperinflammatory and immunosuppressive signatures in monocytes. Cell Rep 2021; 37(1): 109793.

14. Mann ER, Menon M, Knight SB, et al. Longitudinal immune profiling reveals key myeloid signatures associated with COVID-19. Sci Immunol 2020; 5(51).

15. Kreuter M, Lee JS, Tzouvelekis A, et al. Monocyte Count as a Prognostic Biomarker in Patients with Idiopathic Pulmonary Fibrosis. Am J Respir Crit Care Med 2021; 204(1): 74–81.

16. Scott MKD, Quinn K, Li Q, et al. Increased monocyte count as a cellular biomarker for poor outcomes in fibrotic diseases: a retrospective, multicentre cohort study. Lancet Respir Med 2019; 7(6): 497–508.

17. Szabo PA, Dogra P, Gray JI, et al. Longitudinal profiling of respiratory and systemic immune responses reveals myeloid cell-driven lung inflammation in severe COVID-19. Immunity 2021; 54(4): 797–814 e6.

18. Marais C, Claude C, Semaan N, et al. Myeloid phenotypes in severe COVID-19 predict secondary infection and mortality: a pilot study. Ann Intensive Care 2021; 11(1): 111.

19. Utrero-Rico A, Gonzalez-Cuadrado C, Chivite-Lacaba M, et al. Alterations in Circulating Monocytes Predict COVID-19 Severity and Include Chromatin Modifications Still Detectable Six Months after Recovery. Biomedicines 2021; 9(9).

20. Ryan FJ, Hope CM, Masavuli MG, et al. Long-term perturbation of the peripheral immune system months after SARS-CoV-2 infection. BMC Med 2022; 20(1): 26.

21. Phetsouphanh C, Darley DR, Wilson DB, et al. Immunological dysfunction persists for 8 months following initial mild-to-moderate SARS-CoV-2 infection. Nat Immunol 2022; 23(2): 210–6.

22. Patterson BK, Francisco EB, Yogendra R, et al. Persistence of SARS CoV-2 S1 Protein in CD16+ Monocytes in Post-Acute Sequelae of COVID-19 (PASC) up to 15 Months Post-Infection. Front Immunol 2021; 12: 746021.

23. Guaraldi G, Milic J, Cesari M, et al. The interplay of post-acute COVID-19 syndrome and aging: a biological, clinical and public health approach. Ageing Res Rev 2022; 81: 101686.

24. Stanojevic S, Kaminsky DA, Miller MR, et al. ERS/ATS technical standard on interpretive strategies for routine lung function tests. Eur Respir J 2022; 60(1).

25. SahBandar IN, Ndhlovu LC, Saiki K, et al. Relationship between Circulating Inflammatory Monocytes and Cardiovascular Disease Measures of Carotid Intimal Thickness. J Atheroscler Thromb 2020; 27(5): 441–8.

26. Affandi AJ, Olesek K, Grabowska J, et al. CD169 Defines Activated CD14(+) Monocytes With Enhanced CD8(+) T Cell Activation Capacity. Front Immunol 2021; 12: 697840.

27. Doehn JM, Tabeling C, Biesen R, et al. CD169/SIGLEC1 is expressed on circulating monocytes in COVID-19 and expression levels are associated with disease severity. Infection 2021; 49(4): 757–62.

28. Hartnell A, Steel J, Turley H, Jones M, Jackson DG, Crocker PR. Characterization of human sialoadhesin, a sialic acid binding receptor expressed by resident and inflammatory macrophage populations. Blood 2001; 97(1): 288–96.

29. York MR, Nagai T, Mangini AJ, Lemaire R, van Seventer JM, Lafyatis R. A macrophage marker, Siglec-1, is increased on circulating monocytes in patients with systemic sclerosis and induced by type I interferons and toll-like receptor agonists. Arthritis Rheum 2007; 56(3): 1010–20.

30. Biesen R, Demir C, Barkhudarova F, et al. Sialic acid-binding Ig-like lectin 1 expression in inflammatory and resident monocytes is a potential biomarker for monitoring disease activity and success of therapy in systemic lupus erythematosus. Arthritis Rheum 2008; 58(4): 1136–45.

31. Xiong YS, Cheng Y, Lin QS, et al. Increased expression of Siglec-1 on peripheral blood monocytes and its role in mononuclear cell reactivity to autoantigen in rheumatoid arthritis. Rheumatology (Oxford) 2014; 53(2): 250–9.

32. Darawshy F, Abu Rmeileh A, Kuint R, et al. Residual symptoms, lung function, and imaging findings in patients recovering from SARS-CoV-2 infection. Intern Emerg Med 2022; 17(5): 1491–501.

33. Abdallah SJ, Voduc N, Corrales-Medina VF, et al. Symptoms, Pulmonary Function, and Functional Capacity Four Months after COVID-19. Ann Am Thorac Soc 2021; 18(11): 1912–7.

34. Smet J, Stylemans D, Hanon S, Ilsen B, Verbanck S, Vanderhelst E. Clinical status and lung function 10 weeks after severe SARS-CoV-2 infection. Respir Med 2021; 176: 106276.

35. Daher A, Balfanz P, Cornelissen C, et al. Follow up of patients with severe coronavirus disease 2019 (COVID-19): Pulmonary and extrapulmonary disease sequelae. Respir Med 2020; 174: 106197.

36. Pan Y, Jiang X, Yang L, et al. SARS-CoV-2-specific immune response in COVID-19 convalescent individuals. Signal Transduct Target Ther 2021; 6(1): 256.

37. Schultheiss C, Willscher E, Paschold L, et al. The IL-1beta, IL-6, and TNF cytokine triad is associated with post-acute sequelae of COVID-19. Cell Rep Med 2022; 3(6): 100663.

38. Pandori WJ, Padgett LE, Alimadadi A, et al. Single-cell immune profiling reveals long-term changes in myeloid cells and identifies a novel subset of CD9(+) monocytes associated with COVID-19 hospitalization. J Leukoc Biol 2022; 112(5): 1053–63.

39. Magesh S, John D, Li WT, et al. Disparities in COVID-19 Outcomes by Race, Ethnicity, and Socioeconomic Status: A Systematic-Review and Meta-analysis. JAMA Netw Open 2021; 4(11): e2134147.

40. Tai DBG, Sia IG, Doubeni CA, Wieland ML. Disproportionate Impact of COVID-19 on Racial and Ethnic Minority Groups in the United States: a 2021 Update. J Racial Ethn Health Disparities 2022; 9(6): 2334–9.

41. Penaia CS, Morey BN, Thomas KB, et al. Disparities in Native Hawaiian and Pacific Islander COVID-19 Mortality: A Community-Driven Data Response. Am J Public Health 2021; 111(S2): S49–S52.

42. Dela Cruz MRI, Glauberman GHR, Buenconsejo-Lum LE, et al. A Report on the Impact of the COVID-19 Pandemic on the Health and Social Welfare of the Filipino Population in Hawai’i. Hawaii J Health Soc Welf 2021; 80(9 Suppl 1): 71–7.

